# Patient-centered Transcriptomic and Multimodal Neuroimaging Determinants of Clinical Progression, Physical Activity and Treatment Needs in Parkinson’s Disease

**DOI:** 10.1101/2024.09.25.24314374

**Authors:** Quadri Adewale, Ahmed Faraz Khan, Sue-Jin Lin, Tobias R. Baumeister, Yashar Zeighami, Felix Carbonell, Daniel Ferreira, Yasser Iturria-Medina

## Abstract

Parkinson’s disease is a complex and multifactorial disorder, but how its biological and clinical complexity emerge from molecular to macroscopic brain interactions remains poorly understood. Here, we use a personalized multiscale generative brain model to characterize direct spatiotemporal links between genes and multimodal neuroimaging-derived biological factors in PD. We identified a set of genes modulating PD-associated longitudinal changes in dopamine transporter level, neuronal activity, dendrite density and tissue atrophy. Inter-individual heterogeneity in the gene-mediated biological mechanisms is associated with five distinct configurations of PD motor and non-motor symptoms. Although characterized by distinctive biological pathways, all the symptom configurations are associated with cell cycle processes. Notably, the protein-protein interaction networks underlying these configurations revealed distinct hub genes including *MYC, CCNA2, CCDK1, SRC, STAT3* and *PSMD4*. We also uncovered the biological mechanisms associated with physical activities performance in PD, and observed that leisure and work activities are principally related to neurotypical cholesterol homeostasis and inflammatory response processes, respectively. Finally, patient-tailored *in silico* gene perturbations revealed a set of putative disease-modifying drugs with potential to effectively treat PD, most of which are associated with dopamine reuptake and anti-inflammation. Our study constitutes the first self-contained multiscale approach providing comprehensive insights into the complex multifactorial pathogenesis of PD, unravelling key biological modulators of physical and clinical deterioration, and serving as a blueprint for optimum drug selection at personalized level.

## INTRODUCTION

Parkinson’s disease (PD) is a pervasive neurodegenerative disorder that presents with a variety of clinical manifestations such as motor (e.g., rigidity, resting tremor, bradykinesia), psycho-cognitive (e.g., cognitive decline, depression, anxiety) and autonomic symptoms (e.g., constipation, hyposmia, sleep disorder). However, patients display heterogeneous combinations of symptoms, severity, and disease progression. Therefore, the etiopathogenesis of PD points to multiple probable causes including genetics, environment, and lifestyle (Simon, Tanner, and Brundin 2020). But the complex interplay between these biological factors is not clearly understood. Moreover, the pathological processes leading to the disease recruit many biological pathways at different cellular and molecular levels (Dong-Chen et al. 2023). Thus, a comprehensive framework incorporating several disease-associated variables is crucial for advancing the understanding of the disease. This is further supported by the recent efforts in transitioning towards a biological definition of PD (Simuni et al. 2024; Höglinger et al. 2024)

Current PD treatments are only symptomatic, and no single drug addresses the wide range of symptoms seen in patients. The mainstay of PD treatment, dopamine replacement therapy, relieves motor symptoms for a considerable number of patients, especially at the early stage of disease (Armstrong and Okun 2020). However, 9-16% of patients do not respond to dopamine-based therapies, suggesting that patient heterogeneity plays a pivotal role in treatment response. Disappointingly, even hitherto responsive patients subsequently experience medication dose wear off and drug-associated worsening of symptoms, including drug-resistant tremor and medication-induced dyskinesias. As an adjuvant non-pharmacologic intervention, physical therapy helps improve a broad range of symptoms (Armstrong and Okun 2020; Mak et al. 2017). However, the biological mechanisms underpinning the interaction of physical activity with PD are not fully understood. Furthermore, despite the perceived benefits of physical activity, sedentariness is still found among PD patients due to debilitating motor symptoms and other barriers such as perceived low expected benefit, lack of time, fear of falling, etc., that prevent patients from conducting exercise regimens (Ellis et al. 2013). Understanding the biological substrates of physical activity in PD can facilitate the discovery of pharmacological alternatives, especially for those patients in advanced stages of the disease when physical activities are almost impractical.

Neuroimaging measures offer standard tools for routine clinical diagnosis of neurodegenerative diseases, further enabling the elucidation of disease pathogenesis and progression. Initial attempts integrating several neuroimaging modalities with either gene expression or receptor densities provided insights into the multiscale interactions in healthy aging and Alzheimer’s disease (Adewale et al. 2021; Khan et al. 2021). This approach, called multifactorial causal modelling (MCM) (Iturria-Medina et al. 2017), affords a mechanistic way of understanding how the longitudinal changes in a biomarker emerge from the complex interplay between several biomarkers. Unified multimodal neuroimaging and expression of hundreds of genes revealed critical genetic determinants of healthy aging and Alzheimer’s disease, as well as biological mechanisms separating the two processes (Adewale et al. 2021). The applicability of such unified multifactorial approach to subject-level modelling offers the unprecedented opportunity to harness inter-patient heterogeneity for better treatment plans and clinical trial design.

In this study, we extend the multiscale characterization of PD in four fundamental ways: (i) integrating whole-brain gene expression with longitudinal molecular, functional and (micro)structural neuroimaging-derived biological factors to infer gene-mediated brain reorganization in 89 PD patients from the Parkinson’s Progression Markers Initiative (PPMI) cohort, (ii) linking different configurations of PD symptoms to distinct biological mechanisms and protein-protein interaction networks, (iii) identifying molecular mediators of the interplay between PD progression and physical activity, and (iv) using patient-level *in silico* gene perturbations to identify putative disease-modifying drugs for PD. This work represents a pioneering attempt to unify multiple aspects of PD-associated biomarkers and physical activities at different resolutions, paving the way for a deeper understanding of PD biological mechanisms and identifying effective personalized treatments.

## RESULTS

### Whole-Brain Multiscale Transcriptomic-Neuroimaging Model of Parkinson’s Disease

To characterize widespread molecular, functional and structural brain changes in PD patients at the individual level, we fit a whole brain model with gene expression and six longitudinal neuroimaging-derived biological factors. These imaging modalities macroscopically capture typical neurodegenerative changes, namely, dopaminergic loss (DAT-SPECT), neuronal activity (fALFF), directed microstructural changes (fractional anisotropy), undirected microstructural damage (mean diffusivity), dendrite density (t1/t2 ratio (Righart et al. 2017)]), and atrophy (gray matter density). They are acquired over multiple scans in 89 PD patients from the PPMI cohort. The transcriptomic data was derived from 6 neurotypical brains from the Allen Human Brain Atlas (AHBA) (Hawrylycz et al. 2012) across 976 landmark genes, which have been shown to be central to biological functions and recapitulate about 89% of the whole human transcriptome (Subramanian et al. 2017). Anatomical connectivity was estimated from the high-resolution Human Connectome Project template (HCP-1065; *Methods: Anatomical connectivity estimation*).

Our mathematical framework, named gene expression multifactorial causal model (GE- MCM; Figure 1A), is formulated to capture the influence of gene expression on a particular biological factor and accounts for the network-mediated spreading of the subsequent aberrant changes across the brain (see *Methods: Gene Expression Multifactorial Causal Model*). Using a robust Bayesian optimization technique, we estimated regression coefficients (gene-imaging parameters) that capture the modulation effect of each gene on the dynamic changes and interactions of the individual imaging derived biological factor. Even though we used a single fixed neurotypical gene expression template across the population (AHBA), the personalized gene-imaging parameters quantify individual gene dysregulation patterns and serve as proxies for gene-specific deviations needed for individual model fitting. Indeed, when applied to the studied PD population, the model showed a good predictive ability to reproduce the six disease-affected longitudinal imaging-derived biological factors (R^2^ = 0.71±0.2). Notably, the model parameters demonstrated the capacity to (i) correctly unravel the biological mechanisms underlying inter-patient variability in clinical manifestations or physical activity (Figure 1B) and (ii) infer patient-specific complete model for *in silico* drug discovery via gene perturbation (Figure 1C).

**Figure 1:**
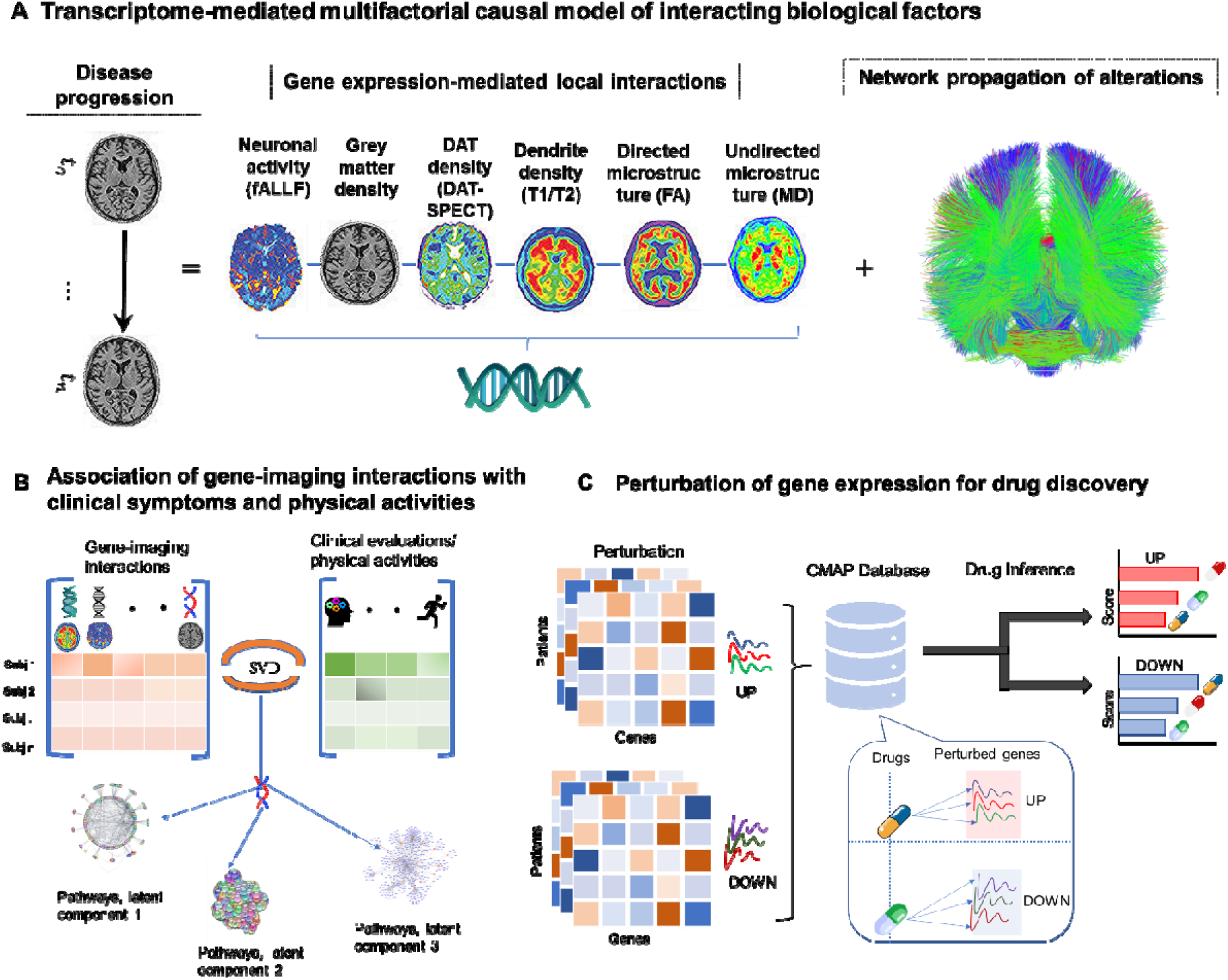
Transcriptomic-neuroimaging multifactorial causal modeling of PD. **A)** Patient’s temporal disease evolution captured by multimodal neuroimaging is decomposed into i) local transcriptome-mediated interactions between neuroimaging measures disease-related biological factors, namely dopaminergic loss, neuronal activity, directed microstructural changes, undirected microstructural damage, dendrite density, and neuronal atrophy ii) network-mediate propagation of pathological effects between brain regions. The patient-specific gene-imaging parameters {α} are obtained by robust Bayesian regressions optimizing the differential equations (*Methods*). **B)** Covariance between the gene-imaging parameters and slopes of clinical evaluations or physical activities are resolved along multiple principal axes to unveil the underlying biological pathways. **C)** *In silico* bidirectional perturbation of genes identifies putative PD drugs. The perturbation of a therapeutic gene is expected to cause a slower disease progression when compared to progression without perturbation.

### Identifying Transcriptomic Mechanisms Mediating Behavioural and Cognitive Deterioration in PD

We sought to identify genetic drivers of multifactorial brain reorganization due to PD progression, particularly those genes controlling direct spatiotemporal interactions among dopaminergic loss, neuronal activity, directed microstructural changes, undirected microstructural damage, dendrite density, and atrophy. First, out of a total of 35,136 gene-imaging parameters, we identified 953 stable parameters whose 95% confidence intervals (CI) exclude zero. Singular value decomposition (SVD) was then used to find the shared latent space between these stable parameters and 11 different clinical evaluations (*Methods: Clinical and Physical Activity Measures*). Five of the eleven principal components are significant following permutation tests (p < 0.05). However, the first principal component accounts for a notable proportion (43.7%) of the explained covariance. Projection of the gene-imaging interactions and clinical evaluations on this first latent component showed a very high correlation of r = 0.93 (p = 0.001: Figure 2A). Furthermore, we discovered 85 genes with significant contributions to the axis (bootstrap ratio > 1.96). Interestingly, querying the diseases associated with the genes in DisGeNET database revealed PD as the leading disorder (q<0.05; Figure 2B). The identification of esophageal carcinoma, medulloblastoma and shigella disease corroborates bodies of evidence associating cancers and gut disorders with PD. Shigella and Escherichia coli are major causes of diarrhea, and Shiga toxins is linked to damage in blood-brain barrier, microvasculature, astrocytes and neuron with characteristic motor symptoms (Pinto et al. 2017). Similarly, α-synuclein (*SNCA*) has been suggested as a biomarker for medulloblastoma (Y.-X. Li et al. 2018). The results support the relevance of the identified genes to PD pathogenesis and its systemic interaction with other disorders.

**Figure 2:**
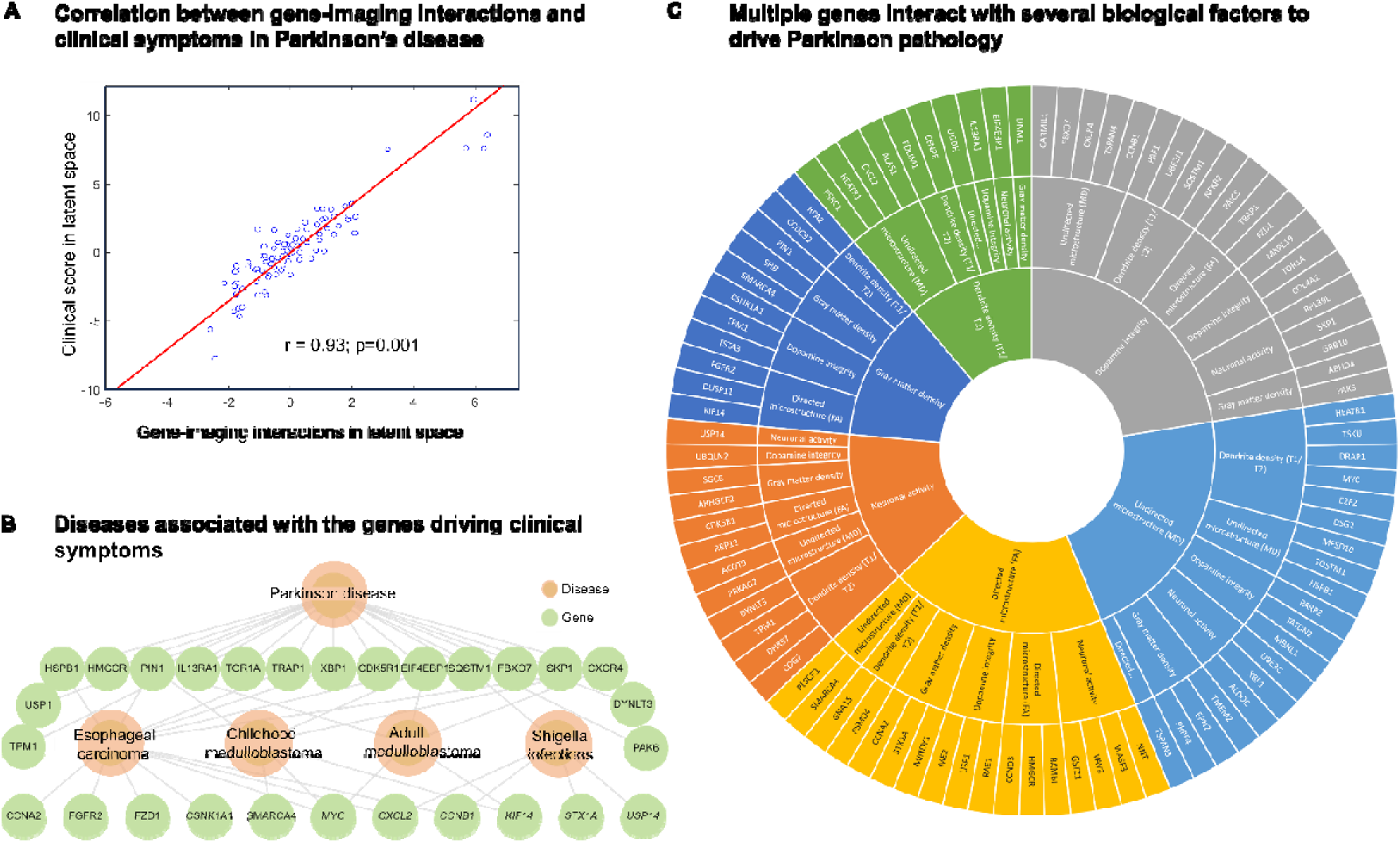
Genes underlying clinically-relevant spatiotemporal vulnerability in PD overlap with cancer and infection pathways. **A)** High correlation (r=0.93; p=0.001, FWE-corrected) between projections of gene-imaging pathological interactions and rates of clinical deterioration on the first latent component of SVD. The first latent component accounts for 43.7% (p=9.99×10^-4^, FWE-corrected) of the explained covariance between clinical evaluations andthe pathological interactions of gene expression and neuroimaging measures . **B)** DisGeNET disease-gene associations of the genes contributing to the pathological interactions on the first latent component. The hierarchical relationship shows the significant cognate diseases and their shared genes (q<0.05). **C)** Multifactorial interactions between identified genes and neuroimaging-derived biological factors. A gene directly influences how a neuroimaging-quantified biological factor interacts with other factors to cause a factorial alteration along the disease’s course. Notably, the outermost ring represents the genes modulating the interactions among biological factors, the middle ring displays the biological factor directly influenced by a gene, and the innermost ring shows the biological factor undergoing longitudinal changes because of the interactions.

We further investigated the specific structural, functional, microstructural, or dopaminergic changes that are modulated by the 85 genes. Since each optimized gene-imaging parameter associates a gene with a biological factor, we retrieved the biological factors of the significant parameters associated with the 85 genes. We observed a broad range of interactions between the genes and the six disease-related biological factors (Figure 2A). Among the PD-related genes, we observed that *TPM1* modulates dopamine level in driving longitudinal changes in atrophy, which is consistent with the gene’s role in controlling striatal dopamine release (Wakabayashi-Ito et al. 2011; Downs et al. 2021). Our results further suggested that *TPM1* also modulates dendrite density to drive longitudinal alterations in neuronal activity, in agreement with the gene’s activity of regulating actin filament and neurite growth (Brettle, Patel, and Fath 2016). Similarly, we found that *CXCR4* modulates mean diffusivity (a measure of myelin or axon integrity) to drive dopamine change longitudinally. Activation of *CXCR4* promotes the development of oligodendrocytes for remyelination of injured adult central nervous system (Patel et al. 2010). Overall, our findings transcend traditional single-scale transcriptomic or neuroimaging analysis by considering biologically plausible complex interactions underlying PD progression.

### Uncovering the Protein-Protein Interaction Networks Underlying PD Phenotypic Landscapes

To understand how the model-derived pathological interactions might be related to the different clinical manifestations of PD, we analysed all the five significant latent components of the SVD. The explained co-variance of these components are 43.7%, 14.5%, 10.2%, 7.1% and 6.3%, respectively. Projecting the 11 clinical scores onto these components allowed us to disentangle the contributions of psychiatric, motor, cognitive and other PD symptoms to each latent component. Using a high confidence score (cut-off=0.7), we then retrieved the protein-protein interaction (PPI) networks of the genes associated with each component from STRING database (Szklarczyk et al. 2021). The biological pathways (q<0.05) relevant to PPI networks were also obtained from Wikipathways.

In contrast to other components, the first latent component, whose genes were earlier associated with PD in Figure 2, shows a balanced contribution from the four groups of symptoms (Figure 3A). This observation indicates that the leading biological mechanisms underlying PD engender a wide range of clinical symptoms. Nevertheless, the largest individual symptomatic contribution comes from motor signs (UPDRS-III), the principal hallmark of PD. The associated PPI network points to the active roles of cell cycle, DNA damage, and rapamycin signaling. The second component is dominated by cognitive symptoms, which supports why the biological pathways include Alzheimer’s disease, in addition to insulin signalling, cell cycle, and gastrin signalling. The largest contributions to the third component come from motor, cognitive and other non-motor symptoms of daily living (e.g., pain, fatigue, and autonomic dysfunctions.). Notably, the implicated pathways include ferroptosis, unfolded protein response, cell cycle and oxidative stress. The fourth and fifth components are predominantly psychiatric and motor symptoms, with suggested roles of inflammation, leptin signalling, cell cycle, DNA damage response, and oxidative stress. Despite the varied symptom profiles and underlying PPI networks, we observe a common association of cell cycle processes with all the latent components.

**Figure 3:**
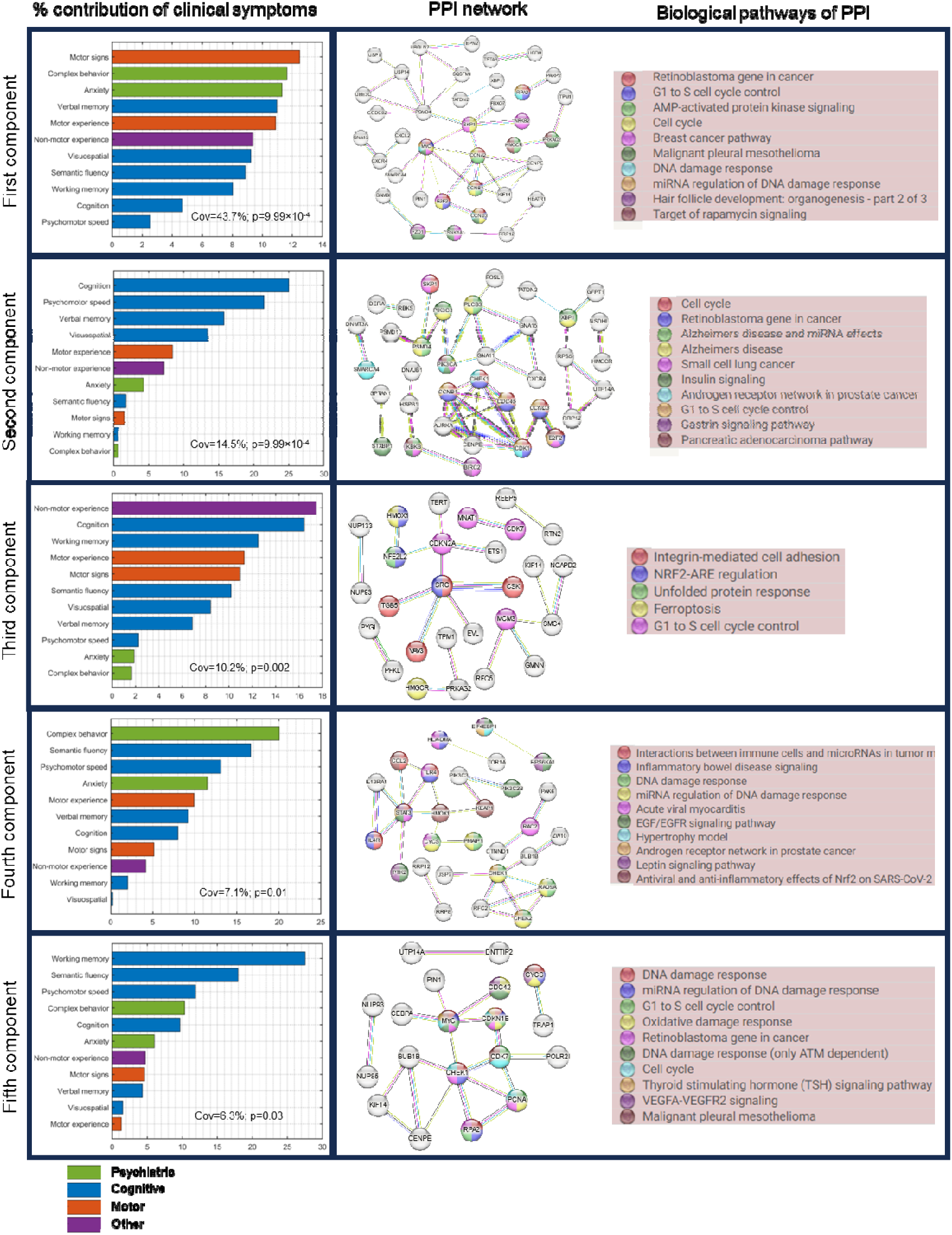
Associations of patterns of PD clinical symptoms with biological mechanisms. *Left:* Five significant latent components were identified based on permutation analysis (FWER<0.05) of shared covariance between gene-imaging pathological interactions and clinical evaluations. Bar graph shows the relative contributions of each clinical evaluations, grouped by symptom types, on each latent component. *Middle:* Protein-protein interaction networks (PPI) of significant genes associated (bootstrap ratio>1.96) with each latent component. PPPI networks were retrieved from STRING with a high confidence score ≥ 0.7. *Right:* Top biological pathways (q<0.05) associated with the genes in the PPI networks. Colored nodes in the PPI networks correspond to genes implicated in the top biological pathways.

As hub genes are believed to play central roles in biological processes and gene regulatory networks (Yu et al. 2017), we sought to identify the leading hub genes in the PPI networks. Interestingly, each latent component has at least one dense PPI sub-network which could be prioritized for biomarker or drug discovery. We therefore selected the hub genes as those with the highest node degrees. We identified 3 hubs genes, namely, *MYC, CCNA2* and *PSMD4* in the first component due to a tie in their rankings. *CDK1*, *SRC*, and *STAT3* were ranked highest for the second, third and fourth components, respectively. The PPI network of the of fifth component was not queried because its enrichment PPI value was not significant (p = 0.194; Figure 3). Apart from *PSMD4,* other genes have been previously identified as hub genes in PD. Our results however suggests that different hub genes might be associated with different patterns of clinical symptoms in PD.

### Molecular Pathways Associated with Physical Activity in PD

Physical activity reduces the risk of developing PD and ameliorates s both motor and non-motor PD symptoms (Paul et al. 2019; Amara et al. 2019; Langeskov-Christensen et al. 2024). Conversely, the symptomatology of PD presents many barriers (such as motor dysfunction, cognitive impairment, depression and apathy) to engaging in physical activities, (Amara et al. 2019). Molecular pathways modulating the relationship between PD and physical activity may therefore shed light onto key neuroprotective mechanisms. We therefore investigated possible biological mechanisms associated with physical activity in PD by applying SVD to identify axes of covariance between the stable gene-neuroimaging parameters and three different domains of physical activity, namely household, work, and leisure activities. The individual scores for the different domains were derived from PASE, a self-reported questionnaire commonly used to quantify physical activity levels in older adults (Washburn et al. 1999). Two SVD principal components were relevant based on permutation tests (p<0.05), and they separately explained 47% and 37% of the data covariance. Leisure activities (e.g., resistance training, jogging, swimming) account for about half (49%) of the first axis (Figure 4. Conversely, work-related activities (e.g., walking and lifting) contribute (54%) principally to the second axis (Figures 4C). Nevertheless, household activity account for 27% and 40% of first and second axes, respectively.

**Figure 4:**
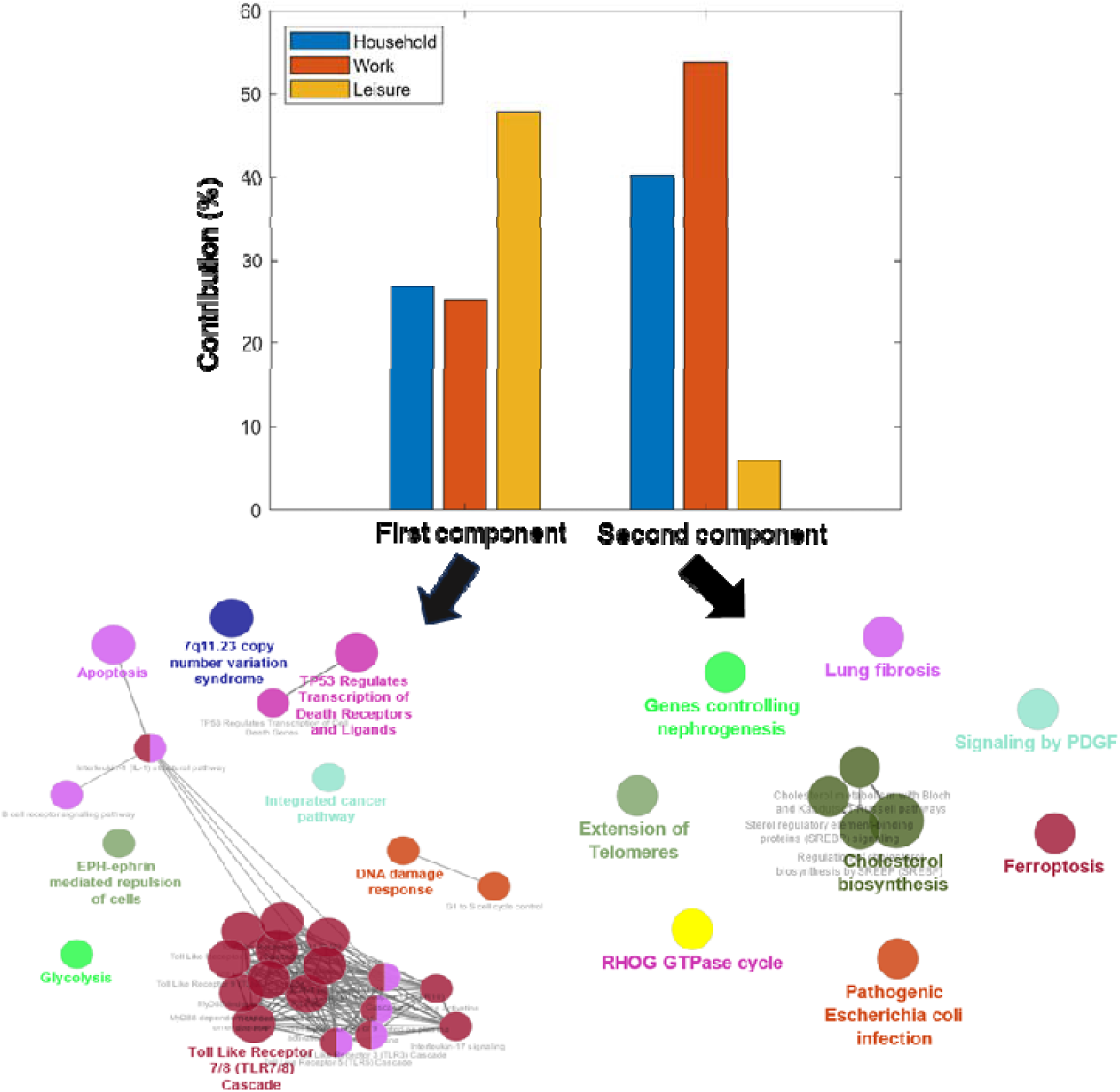
Distinct molecular pathways are associated with leisure and work activities. Contributions of the three domains of the Physical Activity Scale for the Elderly (PASE) towards the first and second principal axes. The corresponding biological pathways mediating the interactions between physical activity and PD in each axis are pointed by the arrows. The primary and secondary axes were obtained from the singular value decomposition (SVD) of the covariance matrix of the gene-neuroimaging parameters and slopes of physical activity scores. The relevant biological pathways were queried from Reactome and Wikipathways (q<0.05)

Next, using the genes with significant contributions to each axis (bootstrap ratio>1.96), we queried the associated biological pathways from Reactome and WikiPathways. The two pathway databases were combined to ensure the robustness of our findings and avoid database bias. The first component is principally associated with cholesterol biosynthesis (q<0.05) while the second component is largely implicated in immune-related processes such as toll-like receptor and B cell signalling. Even though reverse causation cannot be disregarded (as reduced activity levels may accelerate PD progression, and vice versa), the identified biological pathways may be partly explaining individual predisposition/variation to physical activity under PD effects.

### Virtual Gene Perturbations Reveal Potentially Effective Drug Candidates

Finally, we used a multifactorial-causal perspective to predict potential therapeutic drugs candidates for PD. For this, we utilized the individually fitted GE-MCM to simulate the disease’s subsequently progression for 2 years after the last evaluated time point. We then up- and down-perturbed each gene and quantified the influence of each perturbation on the brain’s multiregional and multifactorial imaging descriptors associated with disease progression (*Methods: Gene Perturbation for Drug Discovery*). A gene was considered therapeutic if the perturbation-induced brain changes implied a slower disease progression than the actually observed within the two years under consideration. We then ranked the genes based on the number of subjects for which they have a therapeutic effect and selected the top genes in the 90^th^ percentile. Next, using the CMap database in EnrichR, we queried the inverse-drug relationship between selected genes and several drugs. CMap allowed us to map previous drug-induced transcriptomic perturbations to our *in silico* perturbation profiles. We checked for the alignment between the genes up- and down-regulated by drugs in CMap and our up- and down-perturbed therapeutic genes, respectively. We then retrieved the associated disease and pharmacological classification of these drugs from PubChem.

Figures 5A and 5B show the list of the top respective drug candidates ranked by combined score (product of odds ratio and negative natural log of the p-value). Three of the drugs are associated with dopamine, the principal neurotransmitter implicated in PD. The first among the list of drugs associated with the upwardly perturbed genes is nomifensine, a drug that increases synaptic dopamine availability by inhibiting dopamine reuptake (Figure 5A). Similarly, the fourth drug is Levodopa, the most commonly used drug for treating PD symptoms. Among the top drugs identified through the *in silico* down-perturbation is pergolide, an ergoline-based dopamine receptor agonist still being used to treat PD in some countries. Furthermore, we found a notable number of drugs currently used to treat infections, hinting at the potential of repurposing anti-infectives for PD treatment. Other drugs are implicated in cardiovascular disease, insomnia and inflammation. In sum, the identification of some of the current dopamine-base PD drugs demonstrates the potential of our *in silico* perturbation method to discover prospective drugs for PD treatment.

**Figure 5:**
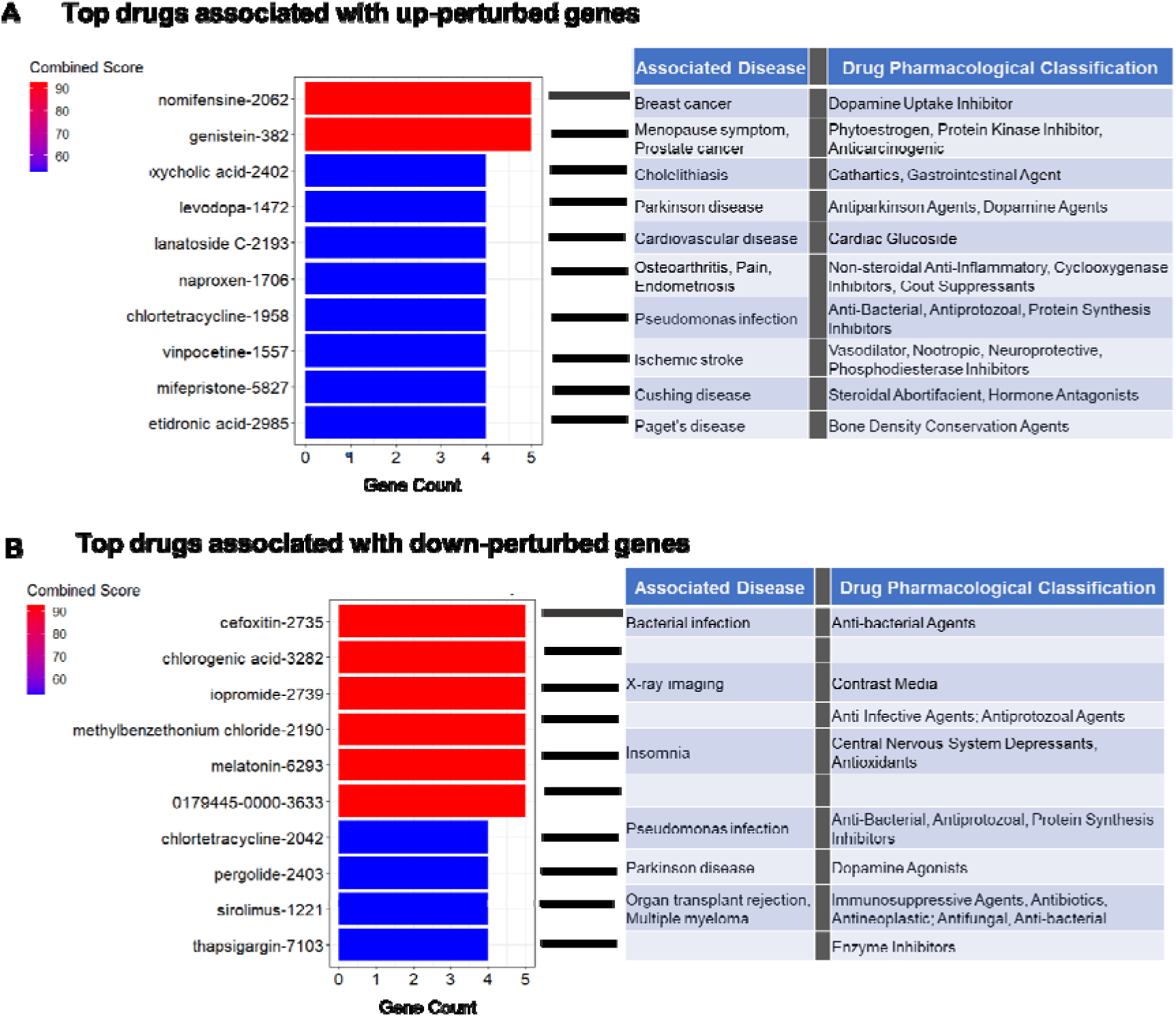
Top drugs for PD identified by our virtual perturbation framework. A) The top 10 drugs identified by upward perturbation of genes. The therapeutic genes were discovered by increasing their expressions by 10% and observing the effect of disease progression within 2 years. The drugs were obtained by comparing the identified therapeutic genes with the transcriptomic effect of drugs from CMAP database. The drugs are ranked by the combined score (odds ratio × -log(p-value)). B) The top 10 drugs identified by downward perturbation of genes.

## DISCUSSION

Parkinson’s disease (PD) is a highly complex and heterogenous disease involving various biological mechanisms. We developed a novel computational approach that incorporates multimodal neuroimaging data, averaged template of bulk gene expression, clinical evaluations, and physical activities to unravel the multifactorial changes accompanying the disease process. We validated the relevance and informativeness of the personalized models by identifying PD as the primary neurodegenerative disease associated with the molecular pathways and clinical symptoms. We further unravelled the biological substrates underpinning the relevance of physical activity to the disease course. Finally, we demonstrated the usefulness of our approach for drug discovery and repurposing via *in silico* transcriptomic perturbations. This first of its kind study presents a self-contained bottom-up causal approach for advancing the understanding of complex multilevel disease processes and identifying potential disease-modifying therapeutic targets.

The roles of genes in maintaining healthy aging and contributing to neurodegenerative diseases are not completely known. In this work, we modelled mechanistic interactions between imaging-derived longitudinal biological factors and spatial variability in gene expression. This modelling approach allowed us to uncover PD-relevant genes and the biological processes they interact with. For instance, among the genes identified are *PIN1*, *SKP1*, *TRAP1, TOR1A* (Figure 2A). *PIN1* is expressed in neurons and found play active roles in neuronal cell death and apoptosis (Ghosh et al. 2013; Zhang et al. 2022). Mice lacking *PIN1* displayed neuronal degeneration including motor and behavioral dysfunctions (Liou et al. 2003). Concordantly, our result suggests that *PIN1* directly modulates gray matter density in PD (Figure 2A). Similarly, decreased expression of *SKP1* homolog (*SKPA*) and *TRAP1* have been shown to cause loss of dopamine in flies, accompanied by motor symptoms (Dabool et al. 2020; Butler et al. 2012). We found that the two genes directly interact with dopamine to drive longitudinal change in neuronal activity and directed microstructure, respectively. *TOR1A* is highly expressed in the substantia nigra, a key region in the pathogenesis of PD, and is responsible for primary hereditary form of dystonia, partly due to its effect on striatal dopamine (Wakabayashi-Ito et al. 2011). Concordantly, our findings suggest that *TOR1A* modulates dopamine to drive the longitudinal alterations in dopamine integrity. Overall, many of the gene-imaging relationships identified in our study of PD patients have also been reported *in vivo* in animal models. Hence, the novel gene- neuroimaging associations can be further validated through experimental models. The insights afforded by these relationships can advance our mechanistic understanding of the disease and help to streamline the identification of possible off-targets when targeting genes for drug development.

Although the hallmark signs of PD are motor complications, dysregulation of multiple clinical domains including cognition, memory, mood, behavior, and autonomic functions supports the complex and multisystem view of the disease. Moreover, heterogeneity in patients’ symptoms and response to treatment has led to the definition of various PD subtypes (Mestre et al. 2021). Even though our study did not subtype patients because of the small sample size, we uncovered five distinct axes of association between biological mechanisms of PD and clinical symptoms. Interestingly, we observed qualitive differences in the relative involvement of symptom types to these axes. Network biology approach also revealed different PPI topology and biological pathways underlying these symptom distribution profiles. Despite the association of numerous pathological processes such as protein aggregation, oxidative stress, ferroptosis, and neuroinflammation with PD, the link between these processes and heterogenous symptom manifestations are lacking. Our study aligns symptom profiles with biological pathways. We found that inflammation, leptin signalling DNA damage response and oxidative stress may be associated with predominant motor and psychiatric symptoms while insulin and gastrin signalling could be implicated in pronounced cognitive symptoms. Nevertheless, we observed a general involvement of G1/S cell cycle control (or its associated processes) with all the symptom distributions. Cell cycle re-entry in post-mitotic neurons might cause neurodegeneration by triggering response to oxidative stress, DNA damage, and other pathological processes (R. Sharma et al. 2017). Concordantly, rotenone-based model of PD showed that lowering the amount of rotenone reduces endoreplication-induced neurodegeneration by blocking cell cycle progression at G1/S phase (H. Wang et al. 2014; Frade and López-Sánchez 2010). We further prioritized 6 hub genes related to these symptom profiles. Interestingly, five of these genes (*MYC*, *CCNA2*, *CCDK1*, *SRC* and *STAT3*) have been previously identified as PD hub genes from different cohort studies of gene expression in substantia nigra and peripheral blood (George et al. 2019; Elango et al. 2023; Liu et al. 2019; M. Wang et al. 2023; Banerjee et al. 2021). The novel hub gene, *PSMD4*, is a receptor of the 26S proteosome which is responsible for protein degradation (Collins and Goldberg 2017). Given the relevance of proteosome homeostasis to intracellular accumulation of α-synuclein (Bi et al. 2021), this novel hub gene may play a key role in PD pathogenesis. The hub genes in this study can guide the identification of druggable targets and biomarkers for heterogenous PD symptom profiles.

The benefits of physical activity to PD symptoms and progression are widely acknowledged. Even though the biological mechanisms mediating these benefits are fully understood, physical activity may promote neuronal plasticity and survival of dopaminergic neurons by simulating the expression of neural growth factors (Da Silva et al. 2016). Here, we found that physical activity is associated with PD through two principal pathways, namely, cholesterol biosynthesis and inflammation via toll-like receptors. A previous study of animal model of PD showed that MPTP-bearing mouse had reduced α-synuclein and downregulation of toll-like receptors after eight weeks of treadmill exercise (Koo et al. 2017). Although the results on the association of cholesterol with PD are mixed, several PD-related genes are involved in cholesterol homeostasis (Jin, Park, and Park 2019; García-Sanz, M.F.G. Aerts, and Moratalla 2021). Moreover, cholesterol biosynthesis has been shown to decrease in the fibroblasts of PD patients (Musanti et al. 1993). The most compelling insights into the tripartite association between PD, cholesterol and physical activity was demonstrated recently (Dutta et al. 2022). The authors found that physical activity activates PPARα in the dopaminergic neurons of PD mouse model. Activation of PPRAα alone suppressed the aggregation and spreading of α- synuclein in the mouse. As PPRAα is a transcription factor that regulates the expression of genes involved in fatty acid oxidation, the mouse was treated with fenofibrate, a PPRAα medication for abnormal cholesterol level. The authors observed that one month of daily treatment with fenofibrate conferred similar benefits as two months of regular exercise. Despite that our analysis does not rule out the bidirectional relationship between PD and physical activity, our results are consistent with the foregoing studies. However, the mode and intensity of exercise remains an open question. A meta-analysis of 19 randomized human clinical trials showed that different modes and regimens of exercise provide different forms of benefits to PD symptoms (Tang, Fang, and Yin 2019). Indeed, our findings could guide a more personalized prescription of physical activity in PD. Perhaps, leisure-related activities (likely shorter duration, higher intensity) would be more beneficial to patients having abnormal cholesterol levels while home- or work-related activities (likely repetitive and lower intensity) could help with neuroinflammation-induced PD pathogenesis. Furthermore, personalized physical activity regimen can be prescribed by comparing the gene- neuroimaging parameters of a patient with the parameters of other patients who have benefited from a particular exercise regimen.

Current treatments for PD are symptomatic, hence the search for disease-modifying treatments addressing the underlying pathology is a priority. While the mainstay of PD treatment are dopamine-based drugs, their effectiveness largely varies with disease subtype and stage (Armstrong and Okun 2020). Interestingly, among the top 20 putative PD drugs identified in our study, there are three dopamine-based drugs, including levodopa, the current first line treatment for PD (Figures 4A and B). However, we also identified multiple immune-related and anti-inflammatory drugs, including naproxen (a non-steroidal anti-inflammatory drug) and tetracycline. Other drugs such as vinpocetine, chlorogenic acid and melatonin have also been reported to modulate inflammation. Although vinpocetine is typically prescribed for treating memory loss in aging and dementias (including PD patients with dementia), it has been demonstrated to regulate the circulation of inflammatory molecules in PD patients (Ping et al. 2019). Chlorogenic acid, found in coffee, is suggested to offer neuroprotective roles in animal models of PD (N. Sharma et al. 2022; Singh et al. 2018; He et al. 2021). This latter finding may partially explain why coffee confers reduced risk on PD development. Similarly, melatonin, which may improve sleep disturbance in PD (Srinivasan et al. 2011), has also been shown to reduce neuroinflammation (Li et al. 2022). The convergence of these medications on immune system/inflammation highlights the need to consider this pathway for drug discovery and repurposing. Even though we performed our drug query using therapeutic genes across the patient population, personalised treatment can be designed by querying the drug database with patient-level therapeutic genes.

The lack of patient-specific gene expression data constrained us to use a single neurotypical gene expression template. Nevertheless, we previously demonstrated that the interaction of the static transcriptomic information with patient-specific longitudinal neuroimaging measures provides a proxy for patient-specific genetic deformation in healthy aging and Alzheimer’s disease (Adewale et al. 2021). Furthermore, the static gene expression data was obtained by combining the mRNA values of six different subjects and inferring the gene expression for the brain regions with missing values (Adewale et al. 2021). Despite the inherent variability and bias that could arise from inter-subject variability and mRNA interpolation, the identification of PD as the underlying neurodegenerative disease demonstrates the validity of our approach (Figure 2B). Subject-specific gene expression may help refine the derived gene-imaging parameters and better facilitate personalized treatments.

Overall, our universal mathematical formulation can be used to study other multifactorial and progressive disorders such as frontotemporal dementias and amyotrophic lateral sclerosis. As subtyping often requires a large number of subjects and raises a question of within-subtype homogeneity, the gene-imaging parameters provide a way to mechanistically capture biological and clinical variability for better treatment plans in heterogenous diseases. Future work will also consider how these parameters can predict patient response to treatment in clinical trials.

## METHODS

### Ethics Statement

This article does not contain any studies with human participants performed by any of the authors. The neuroimaging and clinical data were acquired from the multicenter Parkinson’s Progression Markers Initiative (PPMI; ppmi-info.org). As per PPMI protocols, study participants and/or authorized representatives gave written informed consent at the time of enrollment for sample collection and completed questionnaires approved by each participating site Institutional Review Board (IRB). The authors obtained approval from the PPMI for data use and publication, see documents https://www.ppmi-info.org/documents/ppmi-data-use-agreement.pdf and https://www.ppmi-info.org/documents/ppmi-publication-policy.pdf, respectively.

### Data Description and Processing Study Participants

This study involved 89 individuals from PPMI (RRID:SCR_006431) (http://ppmi-info.org/). The subjects have at least three imaging modalities out of the following: structural MRI, resting functional MRI, diffusion MRI, dopamine SPECT; for at least three visits. Please see Supplementary Table 1 for demographic characteristics. The PPMI was launched in 2010 as an observational study of longitudinal changes in volunteer subjects with and without PD. PPMI is led by Principal Investigator Kenneth Marek, MD and sponsored by the Michael J. Fox Foundation, with the goal of understanding the onset and progression of PD.

### Structural MRI

Structural T1- and T2-weighted 3D brain images were acquired as described in PPMI manuals (http://www.ppmi-info.org/). The images were corrected for intensity nonuniformity using the N3 algorithm (Sled et al., 1998). They were segmented into grey matter (GM), white matter (WM), and cerebrospinal fluid (CSF) probabilistic maps, using SPM12 (http://www.fil.ion.ucl.ac.uk/spm). The gray matter segmentations were standardized to MNI space (Evans et al., 1994) using DARTEL (Ashburner, 2007). Each map was corrected for the effects of spatial registration to preserve the initial amount of tissue volume. Mean gray matter density values of the T1- and T2-weighted images were calculated for a total of 163 grey matter regions described in *Methods: Gene Expression and Brain Parcellation*.

### Resting-State fMRI

Resting-state functional images were acquired using an echo-planar pulse sequence on a 3.0T Philips MRI scanner with the following parameters: 140 time points, repetition time (TR) = 2400 ms, echo time (TE) = 25 ms, flip angle = 80°, number of slices = 40, slice thickness = 3.3 mm, in-plane resolution = 3.3 mm, and in-plane matrix size = 68 × 66. The fMRI images were preprocessesd using FSL (v5.0) toolbox (https://fsl.fmrib.ox.ac.uk/fsl/fslwiki)(S. M. Smith et al. 2004). The preprocessing steps are: 1) Motion and splice timing correction 2) Alignment to the structural T1 image 3) Spatial normalization to the MNI space using the registration parameters obtained for the structural T1 image with the nearest acquisition date, and 4) Signal filtering to retain only low-frequency fluctuations (0.01–0.08 Hz) (Chao-Gan and Yu-Feng, 2010). Due to its high sensitivity to disease progression (Iturria-Medina et al., 2016), we used fractional amplitude of low-frequency fluctuation (fALFF) as a regional quantitative indicator of the brain’s functional integrity fALFF quantifies resting-state regional brain activity as the ratio of the power spectrum of the low frequency band (0.01 – 0.08 Hz) to the power spectrum of the whole frequency range (0 - 0.25Hz) (Zou et al. 2008).

### Diffusion MRI

Diffusion MRI (dMRI) was obtained using standardized protocols on Siemens Verio and Siemens Tim Trio 3T MRI scanners. A single-shot echo-planar imaging scheme was used with 64 sampling directions, a b-value of 1000 s/mm^2^ and a single b = 0 image. Other parameters include 116 × 116 matrix, 2 mm isotropic resolution, TR/TE 900/88 ms, and two-fold acceleration.. More information on the dMRI acquisition and processing can be found online at http://www.ppmi-info.org/. Further preprocessing was done in FSL (v5.0). First, the DTI scans were corrected for motion, eddy current and EPI distortion. Then, the b0 images were aligned to the corresponding subject’s T1-weighted images based on mutual information. The deformation field between the diffusion and T1-weighted image was calculated. The deformation field and eddy current transformations were applied to the dMRI images. Diffusion tensor models were then fitted independently for each voxel Next, the scans were normalized to MNI space (Evans et al. 1994) using the registration parameters obtained for the structural T1 image with the nearest acquisition date. The mean values of the fractional anisotropy and mean diffusivity were estimated for each of the 163 brain regions of interest.

### Dopamine SPECT

A 111-185 MBq (3-5 mCi) bolus injection of I-123 FB-CIT was administered to each participant and the SPECT scans were obtained 4 hours post-injection. Raw projection data was acquired as a 128x128 matrix, after which the SPECT image was reconstructed. The images were preprocessed using SPM12. The scans underwent for attenuation correction and noise reduction using Gaussian blurring with a 3D 6mm filter were applied. The reconstructed and corrected SPECT images were normalized to MNI space (Evans et al. 1994), and average values were calculated for the 163 brain regions of interest.

#### Gene Expression and Brain Parcellation

Microarray data was downloaded from the Allen Human Brain Atlas (AHBA) (RRID:SCR_007416) website (http://www.brain-map.org) (Hawrylycz et al., 2012). The AHBA data consists of mRNA expression in 3702 tissue samples obtained from six l adult human brains, with no known neuropathological history. The data was preprocessed by the Allen Institute to reduce the effects of bias due to batch effects. Description of the processing steps can be found in the technical white paper on AHBA website. For each brain, there are 58,692 probes representing 20,267 unique genes. Leveraging the spatial dependence of gene expression patterns, (Gryglewski et al. 2018), Gaussian kernel regression was applied to predict the mRNA intensity in each of the 3702 samples in MNI space using leave-one-out cross-validation. The probe with the highest prediction accuracy (among the multiple probes for a gene) was chosen as the representative probe for that gene. Next, because mRNA values were not available for all the grey matter voxels of the brain, Gaussian kernel regression was again used to predict the GE for the remaining MNI coordinates without mRNA expression intensity. Thus, the whole-brain GE data was obtained for the selected 20,267 probes/genes. As it was infeasible to use these ∼20,000 AHBA genes for modelling, we therefore selected 976 landmark genes (Supplementary Table 1) (Subramanian et al. 2017). These landmark genes are universally informative transcripts with the capacity to cover most of the information in the whole human transcriptome across a diversity of tissue types. The average expression value of each gene was then calculated for the 163 brain regions of interest.

A brain parcellation was derived from a combination of the Jülich, Brodmann, AAL3 and DISTAL atlases. First, structural T1 images of the four atlases were registered to the MNI ICBM152 T1 template using FSL’s FLIRT affine registration tool. Then, the obtained transformations were used to project the corresponding parcellations to the MNI ICBM152 space using nearest neighbour interpolation. The resulting parcellation has 163 gray matter regions of interest which were used to extract the multimodal imaging data, gene expression, and diffusion-based connectivity matrix.

### Anatomical Connectivity Estimation

The connectivity matrix was constructed in DSI Studio (http://dsi-studio.labsolver.org) using a group average template from 1065 subject (Yeh et al. 2018). A multi-shell high-angular-resolution diffusion scheme was used, and the b-values were 990, 1985, and 2980 s/mm^2^. The total number of sampling directions was 270. The in-plane resolution and slice thickness were 1.25 mm. The diffusion data were reconstructed in MNI space using q-space diffeomorphic reconstruction to obtain the spin distribution function (Yeh and Tseng 2011). The sampling length and output resolution were set to 2.5 and 1 mm, respectively. The restricted diffusion was quantified using restricted diffusion imaging and a deterministic fibre tracking algorithm was used (Yeh et al. 2017). Using the brain atlas previously described under *Methods: Gene Expression and Brain Parcellation*, seeding was placed on the whole brain while setting the QA threshold to 0.15. The angular threshold was randomly varied from 15 to 90 degrees and the step size from 0.5 to 1.5 voxels. The fibre trajectories were smoothed by averaging the propagation direction with a percentage of the previous direction, which was randomly selected from 0 to 95%. Tracks with lengths shorter than 30 mm or longer than 300 mm were discarded. A total of 100,000 tracts were calculated, and the connectivity matrix was obtained by using count of the connecting tracks.

### Multimodal Neuroimaging Modalities

After preprocessing the imaging modalities, the data were harmonized using ComBat (Fortin et al. 2017). As each site used the same scanner for all subjects, the harmonization procedure corrected for batch effects. The harmonized neuroimaging modalities were extracted for 6 measures, namely dopamine SPECT values, fALLF, fractional anisotropy, mean diffusivity, T1/T2 ratio, and gray matter density. Subjects having at least three neuroimaging modalities in at least three time points were selected. For these subjects, the modalities missing at each time point having actual individual data were automatically imputed using the trimmed scores regression with internal PCA (Folch-Fortuny, Arteaga, and Ferrer 2016). Ultimately, a total of 89 subjects were included in the study with all the 6 neuroimaging modalities for an average of 4 (±0.5) time points. The average numbers of imputed time points per neuroimaging modality are presented in Supplementary Table 2.

### Clinical and Physical Activity Measures

For general clinical measures, we used eleven scores obtained from the PPMI testing battery, namely the Benton Judgment of Line Orientation Test (BJLOT) (Woodard et al. 1996), Hopkins Verbal Learning Test (HVLT) (Brandt 1991), Letter Number Sequencing (LNS) (Saklofske and Schoenberg 2011), Geriatric Depression Scale (GDS) (Yesavage 1988), Movement Disorders Society – Unified Parkinson’s Disease Rating Scale (MDS-UPDRS) (Goetz et al. 2008) Parts 1 (non-motor aspects of daily living), 2 (motor aspects of daily living), and 3 (motor examination), the Montreal Cognitive Assessment (MoCA) (Nasreddine et al. 2005), semantic fluency (SF), State-Trait Anxiety Inventory for Adults (STAIAD) (Beckler 2010), and Symbol Digit Modalities (SDM) (A. Smith 1973). For the measures of physical activity, we used the three different subscores of the Physical Activity Scale for the Elderly (PASE) (Washburn et al. 1999), with higher scores indicating higher levels of physical activity. The subscores include PASE leisure score, PASE work score, and PASE household score. The methods for deriving all the composite scores are described in the respective PPMI protocols documentation. For each subject, we calculated the rate of change of the scores with respect to the examination date. The slopes of the clinical and physical scores are then used for subsequent analyses.

### Gene Expression Multifactorial Causal Model (GE-MCM)

The GE-MCM models how alterations in different regional neuroimaging-derived biological factors and their interactions are controlled by regional gene expression patterns in the brain (Adewale et al. 2021; Iturria-Medina et al. 2017). Simply, the model is defined by: (i) the influence of each gene on the local direct interactions among all the macroscopic imaging modalities factors, constrained within each brain region, (ii) the potential spreading of macroscopic factor-specific alterations through anatomical and/or vascular networks. (iii) the temporal changes in each macroscopic imaging factor due to (i) and (ii).

In this work, we considered six biological factors namely, brain atrophy, neuronal activity, dopaminergic neuronal loss, dendritic density, and (un)directed measures of white matter integrity. The factors are derived from T1-weighted MR1, resting-state fMR1, DAT-SPECT, T1/T2 ratio, mean diffusivity and fractional anisotropy, respectively. We also considered the regional mRNA patterns of 976 genes. The temporal evolution of the disease-associated process is thus depicted mathematically as:

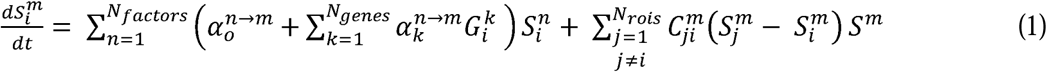

*N_genes_* = 96 is the number of genes normalised across *N_rois_* = 163 brain regions of interest covering most of the brain’s gray matter. Each gene *i* is denoted as *G_i_*, and *N_factors_* =6 is the number of different biological factors measured at the same brain region. The first term on the right-hand side of the equation models the local direct influences of multiple macroscopic biological factors on the given factor *m*. The interaction parameters 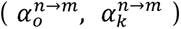 and gene expression 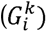 modulate the direct within-region impact of the factor n on m, including intra-factor effects, i.e., when 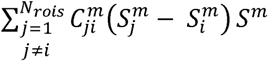 reflects the resultant signal propagation of factor m from region *i* to other brain regions through the physical network 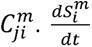 is the local longitudinal alteration of a macroscopic factor *m* at region *i* due to the foregoing multiscale interactions.

### Statistical Analyses

#### Model Fitting

Using the GE-MCM differential equation, for each subject *j* and biological factor *m*, we calculated 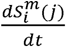 between each pair of consecutive time points. The regional values obtained were concatenated into a subject-factor-specific vector 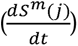 with 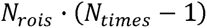 unique values. This allowed us to formulate the identification of the model parameters (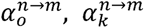) as a regression problem (with 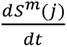 as dependent variable). Due to the high dimensionality of the data, we used a Bayesian sparse linear regression with horseshoe hierarchy to identify the distribution of the model parameters (Carvalho, Polson, and Scott 2010; Makalic and Schmidt 2015). We then obtained regression coefficients (gene-imaging parameters) as a measure of transcriptomic effect on the interaction of a macroscopic imaging-based factor with the other macroscopic factors, in driving a longitudinal biological factor alteration. We calculated coefficient of determination (R^2^) for each subject and neuroimaging modality as a measure of model fit.

#### Covariance of Gene-Neuroimaging Parameters with Clinical Evaluations

Due to high dimensionality, we first reduced the number of gene-imaging parameters by selecting only the stable parameters whose population-wide 95% confidence interval (CI) exclude zero. We then applied singular value decomposition (SVD) to evaluate how the stable transcriptomic-imaging interactions mediate the rates of change in the eleven clinical scores. The aim of SVD is to identify a few pairs of ‘principal components’ that maximize the cross-correlation between the two sets of variables (i.e., gene-imaging interactions and slope of clinical evaluations). We tested for the significance of the identified principal components (PC) by permuting the mapping the gene-imaging parameters and the clinical scores. The permutation was run 1000 times and principal components with a null p<0.05 were considered significant. To identify the genes (gene-imaging parameters) with large and reliable contributions on the significant PCs, we drew 1000 bootstrap samples and calculated the bootstrap ratio of the gene-imaging parameters. The bootstrap ratio is obtained by dividing the saliences (contributions) of the gene-imaging parameters by their respective bootstrap standard errors. Top contributing genes were obtained at a bootstrap ratio>1.96 (corresponding to 95% CI). Diseases associated with the genes were queried from DisGeNET database in Enrichr-KG (Evangelista et al. 2023) at a significance level of q-value<0.05. We derived the PPI networks and the associated WikiPathways from STRING database while setting the PPI confidence score cut-off to 0.7 (Szklarczyk et al. 2021). The hub genes for each PPI network were identified by ranking according to node degrees using cytoHubba plugin (Chin et al. 2014) in Cytoscape (v3.9.1) (Shannon et al. 2003).

#### Covariance of Gene-Neuroimaging Parameters with Physical Activity

We again applied SVD to the stable gene-imaging parameters and the slopes of the three different PASE subscores. Significant principal components were obtained by running 1000 permutation iterations and applying a p-value threshold of 0.05. To identify the top genes mediating physical activity, we drew 1000 bootstrap samples and applied a bootstrap ratio threshold of 1.96 (95% CI). The biological pathways associated with the genes were identified by combining WikiPathways and Reactome databases via the ClueGO (v2.5.9) (Bindea et al. 2009) plugin in Cytoscape. For each of the significant PCs, we evaluated the contribution of each of the PASE subscores by calculating the relative variances along the axis of the PC.

#### Gene Perturbation for Drug Discovery

To discover putative drugs for PD treatment, we sequentially perturbed the gene expressions in both directions. Using Equation (1), gene expression values, most recent neuroimaging measurements, and estimated gene-imaging parameters of each subject, we simulated disease progression for two years, as captured by the longitudinal change of each neuroimaging modality. To perturb a gene, we increased or decreased its expression value by 20% across the population while keeping the values of other genes constant. We then re-simulated the disease progression for 2 years and observed the impact of the perturbation on disease progression. The relative measure of disease progression score is calculated thus:

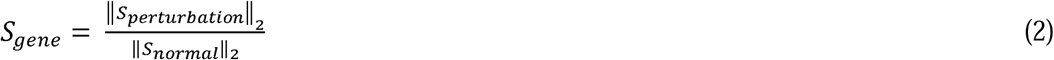

Where *S_gene_* is the relative disease progression score of a gene in a patient. || *S_perturbation_*||_2_ is the norm of 2-year predictions of the six predicted neuroimaging values obtained by perturbing a gene in each direction. || *S_normal_*||_2_ is the norm of 2-year predictions of all neuroimaging modalities without perturbing any gene.

A gene is therapeutic if *S_gene_* < 1; i.e., disease progression induced by perturbation is slower compared to actual disease progression. For each of the perturbation direction, we ranked the genes based on the number of subjects where they are predicted to have therapeutic effects. We selected the genes in the top 90^th^ percentile and used EnrichR (Chen et al. 2013) to query the Connectivity Map (CMap) (Lamb et al. 2006) drug database. Specifically, the top genes that are therapeutic due to upward and downward perturbation are queried using CMap-Up and CMap-Down databases, respectively. Top putative drugs were then ranked by EnrichR combined score. Using PubChem database (Kim et al. 2023), we retrieved the diseases (Therapeutic Target Database (TTD)) and the pharmacological classifications (Medical Subject Headings (MeSH)) associated with the top drugs.

## Data and Code Availability

The three datasets used in this study are available from the PPMI database (neuroimaging and clinical evaluations; https://www.ppmi-info.org/), the HCP database (tractography template for connectivity estimation; http://www.humanconnectomeproject.org/), and Allen Human Brain Atlas website (gene expression; http://human.brain-map.org/static/download). We anticipate that the GE-MCM method will be released soon as part of our available and open-access, user-friendly software (Iturria-Medina et al. 2021) (https://www.neuropm-lab.com/neuropm-box.html).

## Data Availability

The three datasets used in this study are available from the PPMI database (neuroimaging and clinical evaluations; https://www.ppmi-info.org/), the HCP database (tractography template for connectivity estimation; http://www.humanconnectomeproject.org/), and Allen Human Brain Atlas website (gene expression; http://human.brain-map.org/static/download).

## Acknowledgements

The authors thank Dr Konstantinos Poulakis for his advice on statistical analysis. This research was undertaken thanks in part to funding from: the Parkinson Canada and Fonds de recherche du Québec – Santé (FRQS) Graduate Partnership Fellowship awarded to QA, the *Canada First Research Excellence Fund*, awarded to McGill University for the *Healthy Brains for Healthy Lives Initiative*, the Canada Research Chair tier-2, *Fonds de la recherche en santé du Québec* (FRQS) Junior 1 Scholarship, Natural Sciences and Engineering Research Council of Canada (NSERC) Discovery Grant, and Weston Brain Institute awards to YIM, the *Brain Canada Foundation* and *Health Canada* support to the McConnell Brain Imaging Center at the Montreal Neurological Institute, and the *European Union’s Horizon 2020 Framework Programme for Research and Innovation* under the Specific Grant Agreements 785907 (Human Brain Project SGA2) and 945539 (Human Brain Project SGA3) awarded to NPG and KZ. Multimodal imaging and clinical data collection and sharing for this project was funded by PPMI. A public-private partnership, PPMI is funded by the Michael J. Fox Foundation for Parkinson’s Research and funding partners, including AbbVie, Allergan, Amathus Therapeutics, Avid Radiopharmaceuticals, Biogen, BioLegend, Bristol Myers Squibb, Celgene, Denali Therapeutics, GE Healthcare, Genentech, GlaxoSmithKline plc., Golub Capital, Handl Therapeutics, Insitro, Janssen Neuroscience, Eli Lilly and Company, Lundbeck, Merck Sharp & Dohme Corp., Meso Scale Discovery, Neurocrine Biosciences, Pfizer Inc., Piramal Group, Prevail Therapeutics, Roche, Sanofi Genzyme, Servier Laboratories, Takeda Pharmaceutical Company Limited, Teva Pharmaceutical Industries Ltd., UCB, Verily Life Sciences, and Voyager Therapeutics Inc.

## Author Contributions

Q.A. helped conceptualize the project, preprocessed the data, implemented the model, and conducted the analysis. A.F.K, S.-J.L. and Y.Z. helped preprocess the data. T.R.B. and F.C. contributed to statistical methods. Y.I.-M. conceptualized and supervised the project. Q.A. wrote the paper with input from D.F. and Y.I.-M. All authors helped interpret the results and revise the paper.

